# Development of a Transformer-Based Cardiac Arrest Prediction Model for General Ward Patients

**DOI:** 10.64898/2026.01.12.26343973

**Authors:** Sungsoo Hong, Yechan Mun, Kyung Hyun Lee, Sangchul Hahn, Sunguk Jang, Changhun Kim, Kyung Soo Chung, Taeyong Sim

## Abstract

**Background:** In-hospital cardiac arrest on general wards is often preceded by detectable physiological deterioration, yet conventional early warning scores demonstrate limited discrimination. We developed and performed preliminary validation of a transformer-based cardiac arrest prediction system for general ward patients.

**Methods:** This retrospective study was conducted among general ward patients at a tertiary academic hospital in South Korea (Severance Hospital, 2013–2017). We developed Cardiac Arrest Risk Early Detection (CARED), a transformer-based system to predict 24-hour cardiac arrest risk. Model performance was evaluated using the area under the receiver operating characteristic curve (AUROC). Internal validation was performed using 5-fold cross-validation.

**Results:** In internal validation, CARED achieved the highest discrimination with an AUROC of 0.939 (95% CI: 0.928–0.950), significantly outperforming machine learning and deep learning baseline models (all *P* < 0.05)

**Conclusions:** CARED demonstrated high discriminative ability for predicting cardiac arrest in general ward patients. Comprehensive validations including multidimensional performance assessments, comparisons with conventional early warning scores, and external validation in independent populations are ongoing and will be reported in subsequent publications.

## Introduction

In-hospital cardiac arrest (IHCA) affects an estimated 200,000 patients annually in the United States, with survival-to-discharge rates of 15–25% [1,2]. Notably, 59.4% of ward cardiac arrests are preceded by abnormal vital signs in the 1–4 hours before arrest, with increasing abnormalities associated with higher mortality [3]. This window of detectable deterioration presents an opportunity for early intervention; however, identifying at-risk patients remains challenging due to lower monitoring intensity and low event rates in general wards [4,5].

Traditional rule-based early warning scores (MEWS, CART, NEWS) demonstrate limited discrimination (AUROC: 0.60–0.75) and high false-alarm rates [6-9]. Machine learning methods have improved performance (AUROC: 0.78–0.90) [10-13], and deep learning approaches such as LSTM have advanced temporal modeling [14-16]. Transformer architectures further improve long-range dependency capture through self-attention mechanisms [18-20]. However, studies specifically targeting cardiac arrest prediction in general ward patients remain scarce, with most focusing on intensive care, emergency departments, or prehospital settings [21-23].

To address these gaps, we developed Cardiac Arrest Risk Early Detection (CARED), a transformer-based system designed for general ward patients to predict cardiac arrest within 24 hours using multivariate vital sign time series. The objective of this study is to detect subtle deterioration patterns using transformer architecture and to perform comprehensive performance comparisons with traditional machine learning models and baseline deep learning models.

## Methods

### Study Design and Setting

This retrospective study developed a transformer-based early warning system for cardiac arrest prediction at a tertiary academic medical center in South Korea. Development and internal validation utilized patients from Severance Hospital (SH). Institutional Review Board approval was obtained (IRB 4-2017-0939), and patient data were anonymized. We followed TRIPOD+AI reporting guidelines [24].

The study cohort comprised adult patients (≥19 years) admitted to general wards at SH from January 2013 to October 2017. The exclusion criteria were: (1) cardiac arrest in intensive care units, (2) hospitalizations exceeding 30 days, (3) missing measurements of any basic vital signs (systolic blood pressure [SBP], diastolic blood pressure [DBP], pulse rate [PR], respiratory rate [RR], and body temperature [BT]) during general ward stay. A cohort flowchart is in Figure 1.

**Figure 1.**
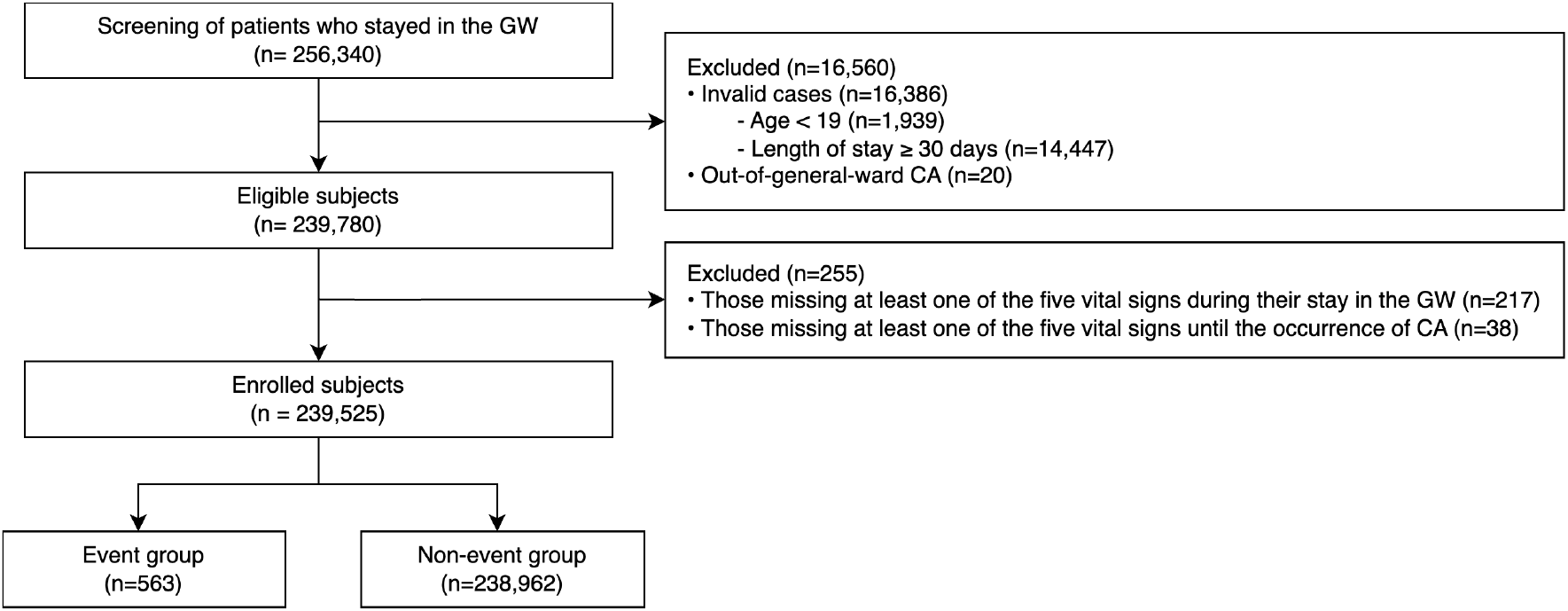
Flowchart of the study population. Abbreviations: CA=Cardiac Arrest; GW=General Ward.

### Outcome and Predictors

The primary outcome was ward cardiac arrest or death in do-not-resuscitate (DNR) patients. For multiple events, the first recorded event was used. The predictors included age and five vital signs (SBP, DBP, PR, RR, and BT). Values identified as outliers or non-numeric entries were treated as missing. Acceptable ranges were: age 18–150 years, SBP and DBP 0–300 mmHg, PR 0–300 beats/min, RR 0–120 breaths/min, and BT 25–50℃.

### Data Preprocessing

The input vectors comprised 48 sequential hourly measurements of five vital signs, with age concatenated separately. Missing measurements within 1-hour intervals were treated as missing data. The risk period was defined as 24 hours preceding outcome occurrence. The input vectors within this period were labeled as events; Otherwise, they were labeled as non-events.

Development and internal validation datasets used different sampling methods. Development included one sample per hospitalization: for negative patients, one random time point from the entire stay; for positive patients, one timepoint from the 24-hour pre-event window. The validation data reflected online monitoring, sampled at all time points with at least one vital sign recorded. For negative patients, entire hospitalization data were used; for positive patients, data from admission to event.

### Model Development

The dataset was split into development (90%) and internal validation (10%) sets through patient-level stratified sampling by outcome to avoid data leakage. Models compared included: logistic regression (LR), random forest (RF), extreme gradient boosting (XGBoost), LSTM, gated recurrent unit with decay (GRU-D) [25], Transformer, bidirectional recurrent imputation for time series (BRITS) [26], and our proposed masked attention-based data embedding (MADE). The MADE model employed a hybrid Transformer-LSTM architecture that applied attention mechanisms and temporal modeling to capture dependencies while handling missing values through masking (Figure 2).

**Figure 2.**
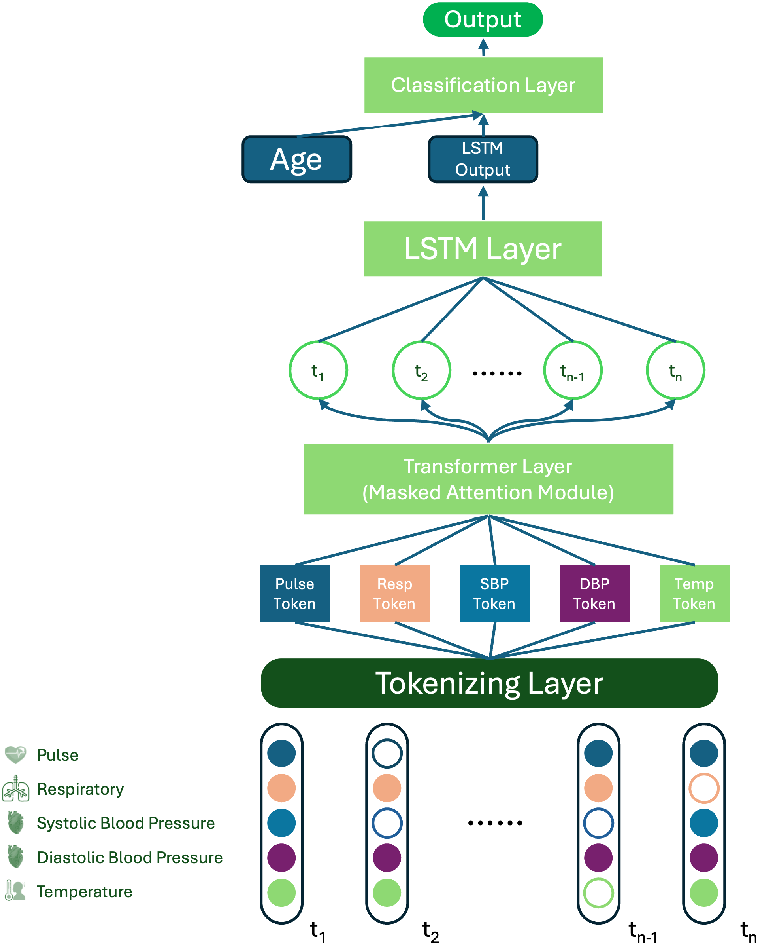
Overview of MADE model architecture. The MADE model integrates a transformer layer with masked attention mechanisms for temporal feature learning from sequential features (e.g., vital signs measured over 48 hours), followed by an LSTM layer for processing. Static patient characteristics (e.g., age) are concatenated with LSTM outputs at the classification layer to generate cardiac arrest risk predictions. The architecture handles missing values through masking without imputation.

For LR, RF, XGBoost, LSTM, GRU-D, BRITS, and Transformer, missing values were forward-filled or imputed with normal values. In contrast, MADE handled missing values through attention masking mechanisms without imputation. Considering class imbalance, random undersampling of non-events was applied during training. Hyperparameters for each model were optimized using a random search, and internal validation was performed using 5-fold cross-validation.

### Performance Evaluation

To evaluate predictive performance, we used the AUROC with 95% confidence intervals (CIs) as the primary metric and the area under the precision-recall curve (AUPRC) for class imbalance. DeLong’s test was used to compare model performance. We also calculated sensitivity, specificity, balanced accuracy (BA), F1 score, positive predictive value (PPV), and negative predictive value (NPV) as additional performance metrics, with 95% CIs estimated through 2000 bootstrap resamples. Optimal thresholds were explored to maximize sensitivity while maintaining specificity above 0.95, reflecting the need to minimize false positive alarms given the low event rate. The highest-performing model was designated as CARED.

### Statistical Analysis

Baseline characteristics were summarized as means with standard deviations (SDs) or medians with interquartile ranges (IQRs) for continuous variables, and counts with percentages for categorical variables. Group differences were assessed using the *t*-test or Mann–Whitney U test for continuous variables and the chi-square test for categorical variables. An absolute standardized mean difference (ASMD) < 0.1 indicated adequate balance [27]. All tests were two-sided; *P*<0.05 was considered statistically significant. All analyses were performed using Python (version 3.12; Python Software Foundation).

## Results

### Baseline Characteristics

The analysis included 239,525 patients admitted to general wards, of whom 563 experienced cardiac arrest (event rate, 0.24%). The cohort was divided into development (n=215,518; event rate, 0.24%) and internal validation (n=24,007; event rate, 0.22%) sets. Baseline characteristics are presented in Table 1. The development and internal validation cohorts were well balanced (ASMD < 0.1). Median age was 60.0 years (IQR 47.0–70.0), and 51.3% were male.

**Table 1.** Baseline characteristics of the development and internal validation cohorts.

| Characteristics | Total<br>(n=239,525) | Development<br>cohort<br>(n=215,518) | Validation<br>cohort<br>(n=24,007) | *P* value | ASMD |
| --- | --- | --- | --- | --- | --- |
| **Sex, n (%)** |  |  |  | <0.001 | 0.033 |
| Female | 116,728 (48.73) | 105,383 (48.90) | 11,345 (47.26) |  |  |
| Male | 122,797 (51.27) | 110,135 (51.10) | 12,662 (52.74) |  |  |
| **Age (years), median**<br>**[IQR]** | 60.00<br>[47.00, 70.00] | 59.00<br>[47.00, 70.00] | 60.00<br>[48.00, 70.00] | <0.001 | 0.038 |
| **Cardiac arrest, n (%)** |  |  |  | 0.786 | 0.002 |
| Yes | 563 (0.24) | 509 (0.24) | 54 (0.22) |  |  |
| No | 238,962 (99.76) | 215,009 (99.76) | 23,953 (99.78) |  |  |
| **Initial vital signs** |  |  |  |  |  |
| Systolic BP, mean<br>(SD) (mmHg) | 127.26 (19.13) | 127.27 (19.13) | 127.15 (19.16) | 0.358 | 0.006 |
| Diastolic BP, mean<br>(SD) (mmHg) | 78.25 (12.10) | 78.26 (12.09) | 78.12 (12.14) | 0.082 | 0.012 |
| Pulse rate, mean (SD)<br>(beats/min) | 80.13 (15.41) | 80.10 (15.38) | 80.45 (15.64) | 0.001 | 0.022 |
| Respiratory rate,<br>median [IQR]<br>(breaths/min) | 20.00<br>[18.00, 20.00] | 20.00<br>[18.00, 20.00] | 20.00<br>[18.00, 20.00] | 0.419 | 0.015 |
| Body temperature<br>median [IQR] (°C) | 36.70<br>[36.50, 37.00] | 36.70<br>[36.50, 37.00] | 36.70<br>[36.50, 37.00] | 0.761 | 0.004 |

Abbreviations: ASMD, absolute standardized mean difference; IQR, interquartile range; SD, standard deviation

### Model Performance

The predictive performances of all models on internal validation are summarized in Figure 3 and Table 2. Among the eight candidate algorithms, MADE showed the highest discriminative ability with an AUROC of 0.939 (95% CI: 0.928–0.950). BRITS and GRU-D demonstrated comparable performance with AUROCs of 0.933 (0.923–0.943) and 0.933 (0.921–0.944), respectively, while XGBoost showed an AUROC of 0.928 (0.916–0.940). LSTM and Transformer achieved AUROCs of 0.918 (0.907–0.929) and 0.913 (0.899–0.926), respectively. In contrast, traditional machine learning models such as LR and RF demonstrated substantially lower AUROC values. DeLong’s test was performed to compare MADE against all other models. MADE significantly outperformed Transformer, LSTM, LR, RF, and XGBoost (all *P* < 0.001). Although BRITS and GRU-D showed high discriminative performance, MADE still showed statistically significant differences (*P* = 0.020 and 0.024, respectively).

**Table 2.**
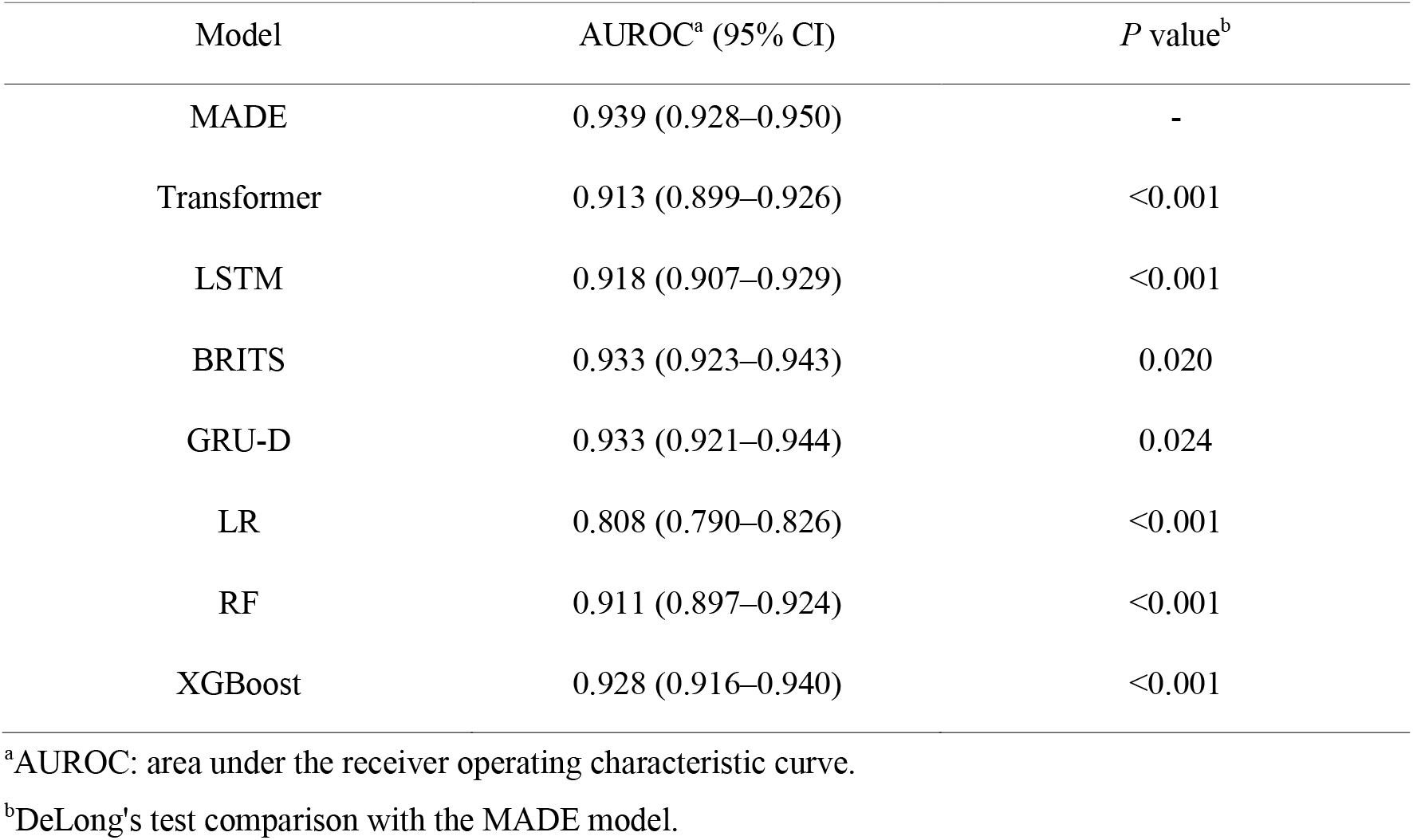
Predictive performance of models in the internal validation.

| Model | AUROC<sup>a</sup> (95% CI) | *P* value<sup>b</sup> |
| --- | --- | --- |
| MADE | 0.939 (0.928–0.950) | - |
| Transformer | 0.913 (0.899–0.926) | <0.001 |
| LSTM | 0.918 (0.907–0.929) | <0.001 |
| BRITS | 0.933 (0.923–0.943) | 0.020 |
| GRU-D | 0.933 (0.921–0.944) | 0.024 |
| LR | 0.808 (0.790–0.826) | <0.001 |
| RF | 0.911 (0.897–0.924) | <0.001 |
| XGBoost | 0.928 (0.916–0.940) | <0.001 |

<sup>a</sup>AUROC: area under the receiver operating characteristic curve.
<sup>b</sup>DeLong's test comparison with the MADE model.

**Figure 3.**
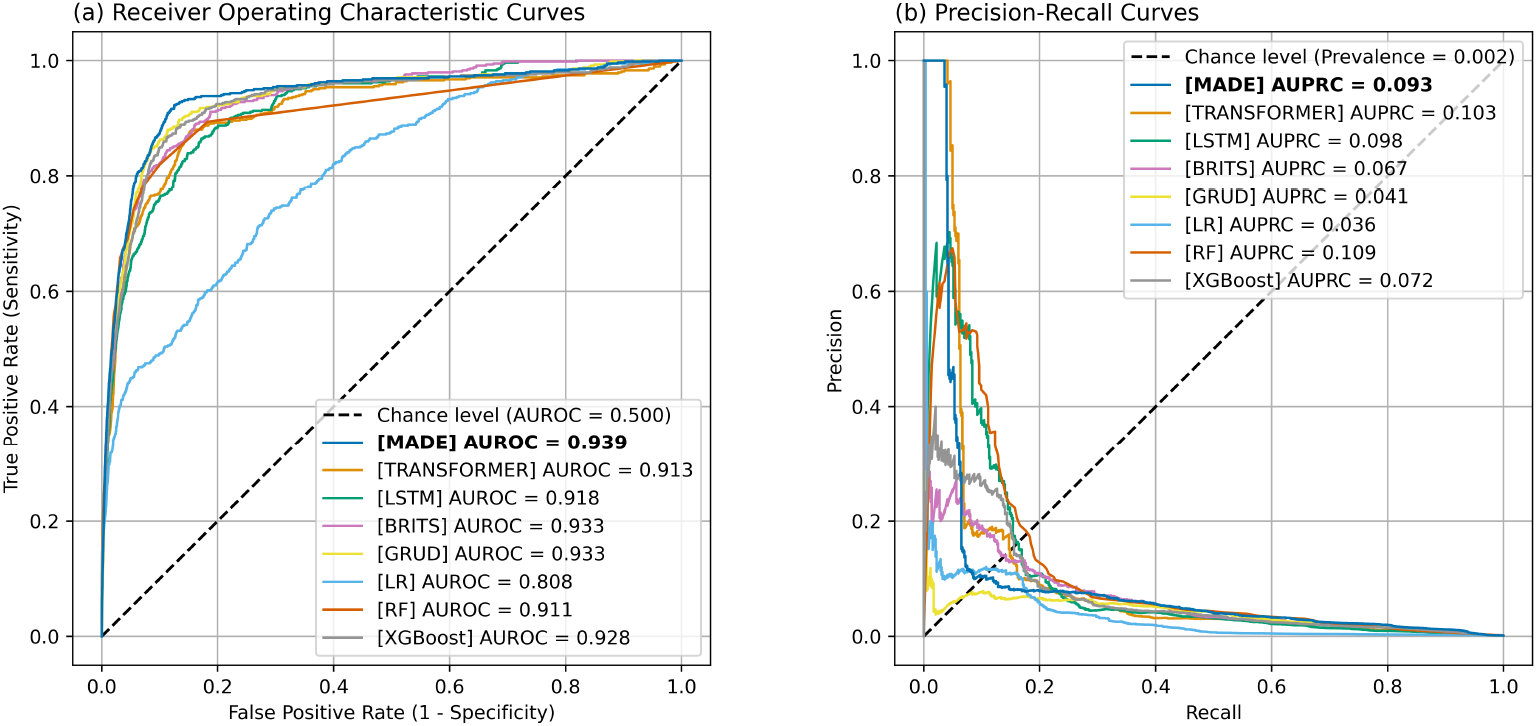
ROC and P-R curves for model performance. The figure compares discriminative performance among candidate algorithms including deep learning models (MADE, Transformer, LSTM, BRITS, and GRU-D) and traditional machine learning models (LR, RF, and XGBoost) in the internal validation.

Given the low event prevalence of 0.22%, AUPRC values ranged between 0.036 and 0.109 across all models. RF demonstrated the highest AUPRC of 0.109 (0.086–0.138), followed by Transformer with 0.103 (0.080–0.128) and LSTM with 0.098 (0.075–0.129). MADE showed an AUPRC of 0.093 (0.074–0.116), while BRITS and XGBoost exhibited AUPRCs of 0.067 (0.054–0.085) and 0.072 (0.057–0.095), respectively. Based on these results, MADE was designated as CARED.

Table 3 shows the performance metrics of CARED at three operating thresholds. At a threshold of 39, determined by maximizing F1 score while maintaining specificity above 0.95, CARED demonstrated a sensitivity of 0.287 (95% CI: 0.250–0.325), specificity of 0.995 (0.994–0.995), PPV of 0.075 (0.065–0.084), and NPV of 0.999 (0.999–0.999). All thresholds maintained a specificity above 99%.

**Table 3.**
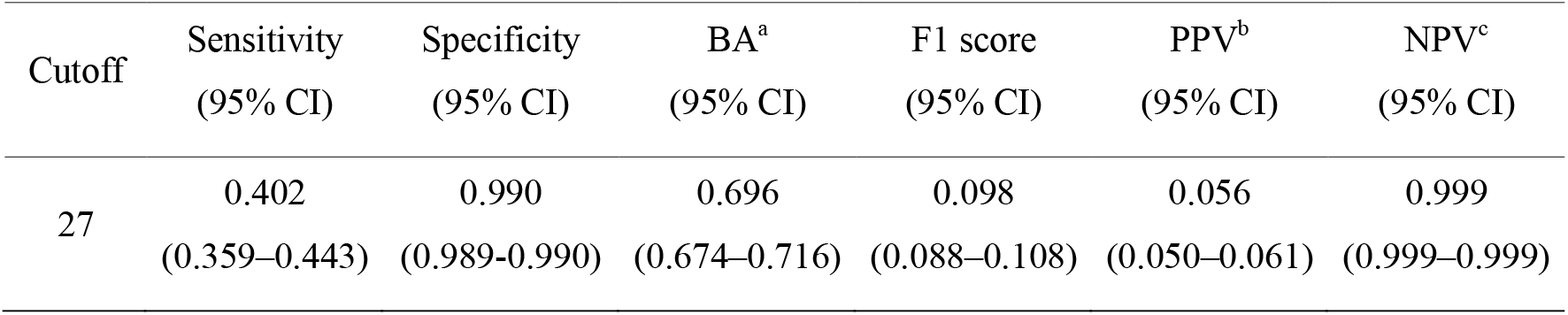
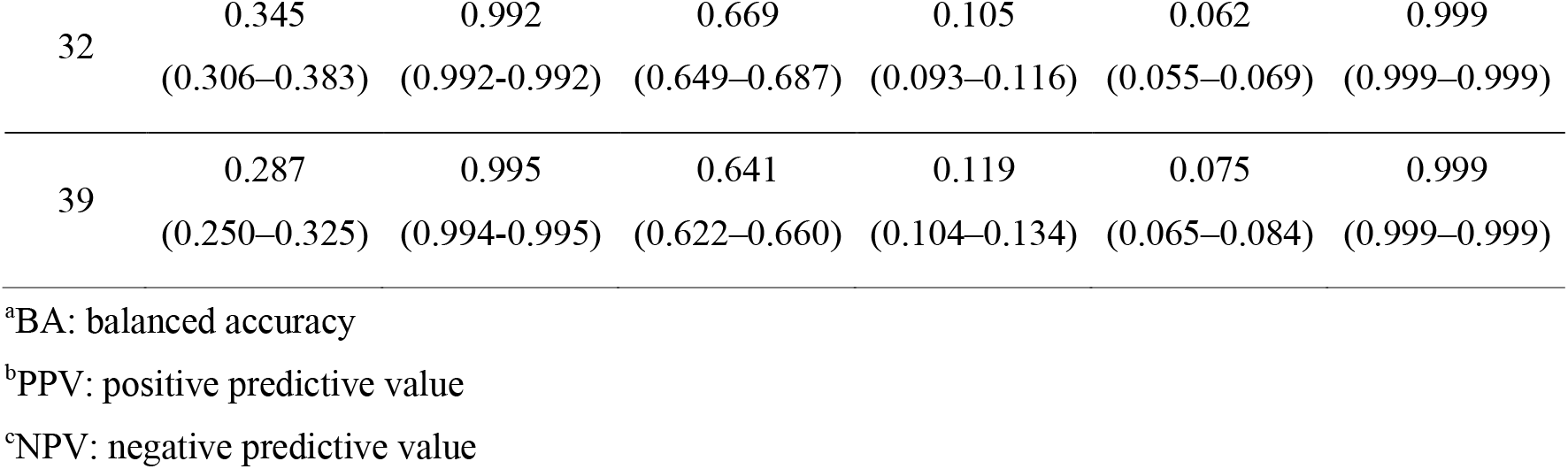
Performance metrics of CARED at three operating thresholds in internal validation. Three operating thresholds were evaluated: a high-sensitivity (cutoff = 27) for comprehensive screening, a balanced (cutoff = 32) for performance comparable to conventional early warning scores, and a high-precision (cutoff = 39) for resource prioritization.

| Cutoff | Sensitivity<br>(95% CI) | Specificity<br>(95% CI) | BA<sup>a</sup><br>(95% CI) | F1 score<br>(95% CI) | PPV<sup>b</sup><br>(95% CI) | NPV<sup>c</sup><br>(95% CI) |
| --- | --- | --- | --- | --- | --- | --- |
| 27 | 0.402<br>(0.359–0.443) | 0.990<br>(0.989–0.990) | 0.696<br>(0.674–0.716) | 0.098<br>(0.088–0.108) | 0.056<br>(0.050–0.061) | 0.999<br>(0.999–0.999) |
| 32 | 0.345<br>(0.306–0.383) | 0.992<br>(0.992–0.992) | 0.669<br>(0.649–0.687) | 0.105<br>(0.093–0.116) | 0.062<br>(0.055–0.069) | 0.999<br>(0.999–0.999) |
| 39 | 0.287<br>(0.250–0.325) | 0.995<br>(0.994–0.995) | 0.641<br>(0.622–0.660) | 0.119<br>(0.104–0.134) | 0.075<br>(0.065–0.084) | 0.999<br>(0.999–0.999) |

<sup>a</sup>BA: balanced accuracy
<sup>b</sup>PPV: positive predictive value
<sup>c</sup>NPV: negative predictive value

## Discussion

### Principal Findings

In this study, we developed a transformer-based early warning system for predicting cardiac arrest within 24 hours in general ward patients using routine vital signs. Among eight candidate algorithms, MADE, designated as CARED demonstrated the highest discriminative ability with an AUROC of 0.939, significantly outperforming traditional machine learning models and deep learning baselines.

### Comparison with the Literature

CARED achieved substantially higher discrimination compared to conventional early warning scores such as MEWS, CART, and NEWS, which typically demonstrate AUROCs of 0.60–0.75. Our results are also consistent with or exceed those of previous machine learning approaches for cardiac arrest prediction, which have reported AUROCs of 0.78–0.90.

The highest performance of CARED may be attributed to the MADE hybrid model’s ability to handle missing values through attention masking without imputation and to capture long-range temporal dependencies. Notably, BRITS and GRU-D, which also employ mechanisms for handling missing data, demonstrated comparable performance to CARED (AUROC of 0.933), suggesting that appropriate treatment of missing values is crucial for clinical time-series prediction in real-world settings where data incompleteness is common.

### Clinical Implications

The high specificity (≥ 99%) maintained across all operating thresholds addresses a critical barrier to clinical adoption, namely alert fatigue. By minimizing false positive alarms while preserving sensitivity, CARED may facilitate earlier recognition of deteriorating patients without overwhelming clinical workflows. The availability of multiple operating thresholds allows clinicians to adjust the system according to institutional resources and clinical priorities, selecting higher sensitivity for comprehensive screening or higher precision for resource-constrained settings. The low event prevalence (0.22%) in our cohort reflects the inherent challenge of cardiac arrest prediction in general wards. Despite this imbalance, CARED maintained robust discrimination, though AUPRC values remained modest across all models, highlighting the difficulty of achieving high precision in rare event prediction.

### Limitations

This study has several limitations. First, we present preliminary internal validation results; more comprehensive evaluations, including multidimensional performance assessments, alarm analyses, and comparisons with conventional early warning scores, are needed. Second, our model was developed using data from a single institution, and external validation in independent populations is required to assess generalizability across different healthcare settings with varying patient populations and monitoring practices. These analyses are ongoing and will be reported in subsequent publications.

## Data Availability

All data produced in the present study are available upon reasonable request to the authors.

## Conclusions

We developed CARED, a transformer-based cardiac arrest prediction system that demonstrated excellent discriminative ability in internal validation. CARED significantly outperformed traditional machine learning and deep learning models, suggesting potential utility as a clinical decision support tool for early detection of deteriorating patients in general wards. External validation and prospective evaluation are warranted to confirm these findings.

## Authors’ Contributions

Sungsoo Hong and Kyung Hyun Lee designed the study, analyzed the data, performed validation, and wrote the manuscript. Yechan Mun developed the predictive models and performed validation. Sunguk Jang and Changhun Kim developed the predictive models. Kyung Soo Chung and Sangchul Hahn collected and preprocessed the data. Taeyong Sim edited the manuscript. All authors reviewed and approved the final version of the manuscript.

## Conflicts of Interest

Sungsoo Hong, Yechan Mun, Kyung Hyun Lee, Sangchul Hahn, Sunguk Jang, Changhun Kim, and Taeyong Sim are employees of AITRICS Co., Ltd., which developed the CARED system. Their roles in this research were limited to those described in the author contributions statement. The remaining authors declare no conflicts of interest.

## Abbreviations

AUROC: area under the receiver operating characteristic curve
AUPRC: area under the precision recall curve
ASMD: absolute standardized mean difference
LR: logistic regression
RF: random forest
XGBoost: extreme gradient boosting
LSTM: long short-term memory
GRU-D: gate recurrent unit with decay
MADE: masked attention-based data embedding
CARED: cardiac arrest risk early detection system

## Notes

### Funding Statement

This study did not receive any funding.

### Author Declarations

This study was approved by the Institutional Review Board of the Yonsei University Health System (4-2017-0939).

## References

1. Holmberg MJ, Ross CE, Fitzmaurice GM, et al. Annual incidence of adult and pediatric in-hospital cardiac arrest in the United States. Circ Cardiovasc Qual Outcomes. 2019;12:e005580.

2. Merchant RM, Yang L, Becker LB, et al. Incidence of treated cardiac arrest in hospitalized patients in the United States. Crit Care Med. 2011;39:2401–6.

3. Andersen LW, Kim WY, Chase M, et al. The prevalence and significance of abnormal vital signs prior to in-hospital cardiac arrest. Resuscitation. 2016;98:112–7.

4. Smith GB. Vital signs: Vital for surviving in-hospital cardiac arrest? Resuscitation. 2016;98:A3–4.

5. Chan PS, Jain R, Nallmothu BK, et al. Rapid response teams: a systematic review and meta-analysis. Arch Intern Med. 2010;170:18–26.

6. Smith GB, Prytherch DR, Meredith P, et al. The ability of the National Early Warning Score (NEWS) to discriminate patients at risk of early cardiac arrest, unanticipated intensive care unit admission, and death. Resuscitation. 2013;84:465–70.

7. Churpek MM, Yuen TC, Winslow C, et al. Multicenter development and validation of a risk stratification tool for ward patients. Am J Respir Crit Care Med. 2014;190:649–55.

8. Alam N, Hobbelink EL, van Tienhoven AJ, et al. The impact of the use of the Early Warning Score (EWS) on patient outcomes: a systematic review. Resuscitation. 2014;85:587–94.

9. Churpek MM, Yuen TC, Park SY, et al. Derivation of a cardiac arrest prediction model using ward vital signs. Crit Care Med. 2012;40(7):2102–2108.

10. Churpek MM, Yuen TC, Winslow C, et al. Multicenter comparison of machine learning methods and conventional regression for predicting clinical deterioration on the wards. Crit Care Med. 2016;44:368–74.

11. Wu TT, Lin XQ, Mu Y, et al. Machine learning for early prediction of in-hospital cardiac arrest in patients with acute coronary syndromes. Clin Cardiol. 2021;44:349–56.

12. Ueno R, Xu L, Uegami W, et al. Value of laboratory results in addition to vital signs in a machine learning algorithm to predict in-hospital cardiac arrest: a single-center retrospective cohort study. PLOS One. 2020;15:e0235835.

13. Yijing L, Wenyu Y, Kang Y, et al. Prediction of cardiac arrest in critically ill patients based on bedside vital signs monitoring. Comput Methods Programs Biomed. 2022;214:106568.

14. Kwon JM, Lee Y, Lee Y, et al. An algorithm based on deep learning for predicting in-hospital cardiac arrest. J Am Heart Assoc. 2018;7:e008678.

15. Lee YJ, Cho KJ, Kwon O, et al. A multicentre validation study of the deep learning-based early warning score for predicting in-hospital cardiac arrest in patients admitted to general wards. Resuscitation. 2021;163:78–85.

16. Baral S, Alsadoon A, Prasad PWC, et al. A novel solution of using deep learning for early prediction cardiac arrest in sepsis patient: enhanced bidirectional long short-term memory (LSTM). Multimed Tools Appl. 2021;80:32639–64.

17. Chen D, Liu S, Kingsbury P, et al. Deep learning and alternative learning strategies for retrospective real-world clinical data. NPJ Digit Med. 2019;2:43.

18. Vaswani A, Shazeer N, Parmar N, et al. Attention is all you need. Adv Neural Inf Process Syst. 2017;30:5998–6008.

19. Li Y, Rao S, Solares JRA, et al. BEHRT: transformer for electronic health records. Sci Rep. 2020;10:7155.

20. Yang Z, Mitra A, Liu W, et al. TransformEHR: transformer-based encoder-decoder generative model to enhance prediction of disease outcomes using electronic health records. Nat Commun. 2023;14:7857.

21. Kang DY, Cho KJ, Kwon O, et al. Artificial intelligence algorithm to predict the need for critical care in prehospital emergency medical services. Scand J Trauma Resusc Emerg Med. 2020;28:17.

22. Fang AHS, Lim WT, Balakrishnan T. Early warning score validation methodologies and performance metrics: a systematic review. BMC Med Inform Decis Mak. 2020;20:111.

23. Hong S, Lee S, Lee J, et al. Prediction of cardiac arrest in the emergency department based on machine learning and sequential characteristics: model development and retrospective clinical validation study. JMIR Med Inform. 2020;8:e15932.

24. Collins GS, Moons KGM, Dhiman P, et al. TRIPOD+AI statement: updated guidance for reporting clinical prediction models that use regression or machine learning methods. BMJ. 2024;385:e078378.

25. Che Z, Purushotham S, Cho K, et al. Recurrent neural networks for multivariate time series with missing values. Sci Rep. 2018;8:6085.

26. Cao W, Wang D, Li J, et al. BRITS: Bidirectional Recurrent Imputation for Time Series. arXiv preprint 1805.10572. 2018.

27. Zhang Z, Kim HJ, Lonjon G, et al. Balance diagnostics after propensity score matching. Ann Transl Med. 2019;7:16.

